# Acupuncture for fatigue in parkinson’s disease: A protocol for systematic review and meta-analysis

**DOI:** 10.1101/2024.02.02.24302182

**Authors:** Yibo Li, Jingxin Zhang, Xiaohan Liu, Tengteng Li, Bingbing Zhang, Xiting Wang, Tao Lu

## Abstract

**Background:** Parkinson’s disease (PD) is a common movement disorder characterized by bradykinesia, rigidity, and resting tremors. Fatigue is a common disabling symptom but is easily ignored in PD. Half of the PD patients were influenced by fatigue. Acupuncture is one of the conservative treatments for fatigue related to other conditions, especially in China. Therefore, we perform a systematic review and meta-analysis to evaluate the evidence for acupuncture’s effectiveness, safety, and cost benefits for the treatment.

**Methods:** This protocol is based on the previously published randomized controlled trial (RCT) studies. A literature search will be performed on the following database: PubMed, the Cochrane Library, Chinese BioMedical Literature Database, China National Knowledge Infrastructure (CNKI), China Science and Technology Journal Database (VIP), and Wanfang Data. According to the Cochrane Risk of Bias Tool and the level of evidence for results, we will assess the quality of the included studies by using the Grading of Recommendations Assessment, Development, and Evaluation (GRADE) method. The Review Manager (v5.3) software will be applied to statistical analysis.

**Results:** From the study, we will assess the effectiveness, safety, and cost-benefit of acupuncture on fatigue relief and functional improvement in patients with Parkinson’s Disease.

**PROSPERO registration number:** CRD42020163155

**Strengths and limitations of this study:** This paper emphasizes the importance of assessing acupuncture in treating Parkinson’s fatigue symptoms and provides methodological guidance for the evaluation of clinical evidence. This study 1) provides a research protocol, 2) facilitates the reasonable evaluation for acupuncture in the treatment of Parkinson’s fatigue symptoms, and 3) raises the potential importance of acupuncture in Parkinson’s fatigue symptoms. Due to the lack of acupuncture reports, there may be a limitation of the small sample size.

## 1. Introduction

Parkinson’s disease (PD) is a common movement disorder characterized by bradykinesia, rigidity, and resting tremors [1]. PD symptoms are divided into two parts: motor symptoms, non-motor symptoms [2]. Non-motor symptoms vary from person to person, and there are several different non-motor symptoms in PD, such as autonomic symptoms and fatigue. Fatigue is a common disabling symptom but is easily ignored in PD [3, 4].

Half of all PD patients were influenced by fatigue [5]. Fatigue in PD may be a clinical manifestation of low-grade systemic hypotension [6]. The study suggests that fatigue is a frequent, independent nonmotor symptom in PD appearing early and persisting throughout the disease course and that establishing uniform diagnostic criteria for PD-related fatigue is critical [7]. At present, Fatigue in PD patients is associated with an increased α-synuclein oligomer level in the cerebrospinal fluid (CSF) [8]. Sang’s study [9] suggests that fatigue in PD may be the expression of metabolic abnormalities and impaired functional interactions between brain regions linked to the salience network and other neural networks.

Roy’s study [10] suggests that rasagiline and modafinil are associated with improved physical fatigue and that doxepin is associated with improved general (combined physical and mental) fatigue. Exergaming [11] was also effective in enhancing balance and reducing fatigue in PD patients after 12 weeks of treatment, but this benefit was not sustained in the long-term. Despite the clear impact of fatigue as a disabling symptom, our understanding of fatigue pathophysiology is limited and current treatment options rarely lead to meaningful improvements in fatigue [12].

It is worth noting that more and more clinical studies have shown that acupuncture has obvious advantages in the adjuvant treatment of PD, such as reducing adverse drug reactions, delaying the course of disease development, improving the quality of life of patients, and improving clinical symptoms [13-15]. In addition, acupuncture can also effectively treat fatigue related to other conditions. However, there is no unified standard for acupuncture treatment of PD in clinical practice, which causes certain obstacles to the rehabilitation and clinical diagnosis andtreatment of PD patients. Therefore, we will make a systematic review and meta-analysis of published RCTs to evaluate the effectiveness and safety of acupuncture for fatigue in PD, to provide more options for clinical external use.

## 2. Materials and Methods

### 2.1 Study registration

The protocol for this systematic review has been registered in PROSPERO (CRD42020163155). Besides, this protocol has been checked with preferred reporting items for systematic review and meta-analysis protocols (PRISMA-P) checklist [16].

### 2.2 Ethics

The approval of an ethics committee is not required for this protocol. Data are not individualized.

### 2.3 Inclusion criteria

#### 2.3.1

Only clinical randomized controlled trials to evaluate the efficacy and safety of acupuncture for fatigue in Parkinson’s disease will be included.

### 2.3.2

Articles posted on PubMed, Google Scholar, the Cochrane Library, Chinese BioMedical Literature Database, China National Knowledge Infrastructure (CNKI), China Science and Technology Journal Database (VIP), and Wanfang Data.

#### 2.3.3

Articles in Chinese and English.

#### 2.3.4

Articles published between January 1970 and May 2022.

#### 2.3.5

Participants who are clinically diagnosed with Parkinson’s Disease with the presence of moderately severe fatigue, aged between 21 and 85, will be included. No restrictions on gender and race.

### 2.4 Exclusion Criteria

#### 2.4.1

Articles in any language other than Chinese and English, published earlier than January 2000, posted on other science websites.

#### 2.4.2

Papers without complete data were not completed even after contacting the authors.

#### 2.4.3

Retrospective or prospective observational studies, including cross-sectional, nested case–control, case–cohort, and cohort studies.

#### 2.4.4

Studies related to acupuncture for Parkinson’s disease but didn’t involve fatigue.

### 2.5 Search strategy

A systematic search will be performed on PubMed, the Cochrane Library, the Chinese BioMedical Literature Database, China National Knowledge Infrastructure (CNKI), China Science and Technology Journal Database (VIP), and Wanfang Data. PubMed MeSH Terms: acupuncture therapy; fatigue; Parkinson disease; acupuncture. Besides, ongoing trials will be retrieved from the WHO ICTRP Search Portal, the Chinese Clinical Trial Register, and The Clinical Trials Register. Details of search strategy are stated as follow: (1) MeSH Terms: acupuncture therapy; fatigue; Parkinson disease; acupuncture; (2) acupuncture; acupuncture therapy; fatigue; Parkinson disease; (3) ((“Acupuncture”[Mesh]) AND “Parkinson Disease”[Mesh]) AND “Fatigue”[Mesh]; (4) ((“Acupuncture”[Mesh]) AND “Acupuncture Therapy”[Mesh]) AND (“Parkinson Disease/ethnology”[Mesh] OR “Parkinson Disease/therapeutic use”[Mesh] OR “Parkinson Disease/therapy”[Mesh])) AND “Fatigue/ethnology”[Mesh] OR “Fatigue/therapy”[Mesh]) . Study titles and abstracts will be screened based on inclusion/exclusion criteria. We will cross-check the references cited as well. We will identify relevant randomized controlled trials from 1970 to May 2022.

### 2.6 Interventions

We will include the studies using acupuncture based on Traditional Chinese Medicine theory as the sole intervention in the experimental group. At the same time, sham needle treatment procedures were utilized in the control group. Studies involving acupuncture combined with other therapies will be included if the other therapies are equally used in both experimental and control groups.

### 2.7 Outcomes

#### 2.7.1 The primary outcome

The primary outcomes are mainly evaluated by (1) the measures of perception of fatigue and subjective fatigue complaints; (2) measures of performance reliability; (3) physiologic factors associated with fatigue or reliability” in Kluger. The related scales can include but are not limited to Modified Fatigue Impact Scale (MFIs), fatigue self-assessment scale (FASA), etc.

#### 2.7.2 The secondary outcome

Motor function, mood, sleep quality, and excessive daytime somnolence, apathy, and quality of life were defined as additional outcomes. Motor function was quantified using the motor subsection of the UPDRS. The mood was assessed using the HADS. Sleep quality and excessive daytime somnolence were assessed using the Parkinson’s Disease Sleep Scale (PDSS) and Epworth Sleepiness Scale (ESS). Apathy was assessed using the Apathy Evaluation Scale (AES). Quality of life (QOL) was assessed using the 39-item Parkinson Disease Questionnaire (PDQ-39).

### 2.8 Data collection and analysis

#### 2.8.1 Study selection

All articles downloaded will be imported into Endnote (X9) to remove the identical studies. According to inclusion and exclusion criteria, two reviewers will independently consult the title and abstract of every searched literature. Any disagreements on study selection will be resolved through discussion with other researchers. According to the Preferred Reporting Items for Systematic Reviews and Meta-Analyses (PRISMA) guidelines[13]. The flow diagram of all study selection procedures is shown in Figure 1. According to the Grading of Recommendations Assessment, Development, and Evaluation (GRADE) method, we will summarize the GRADE judgments.

**Figure 1.**
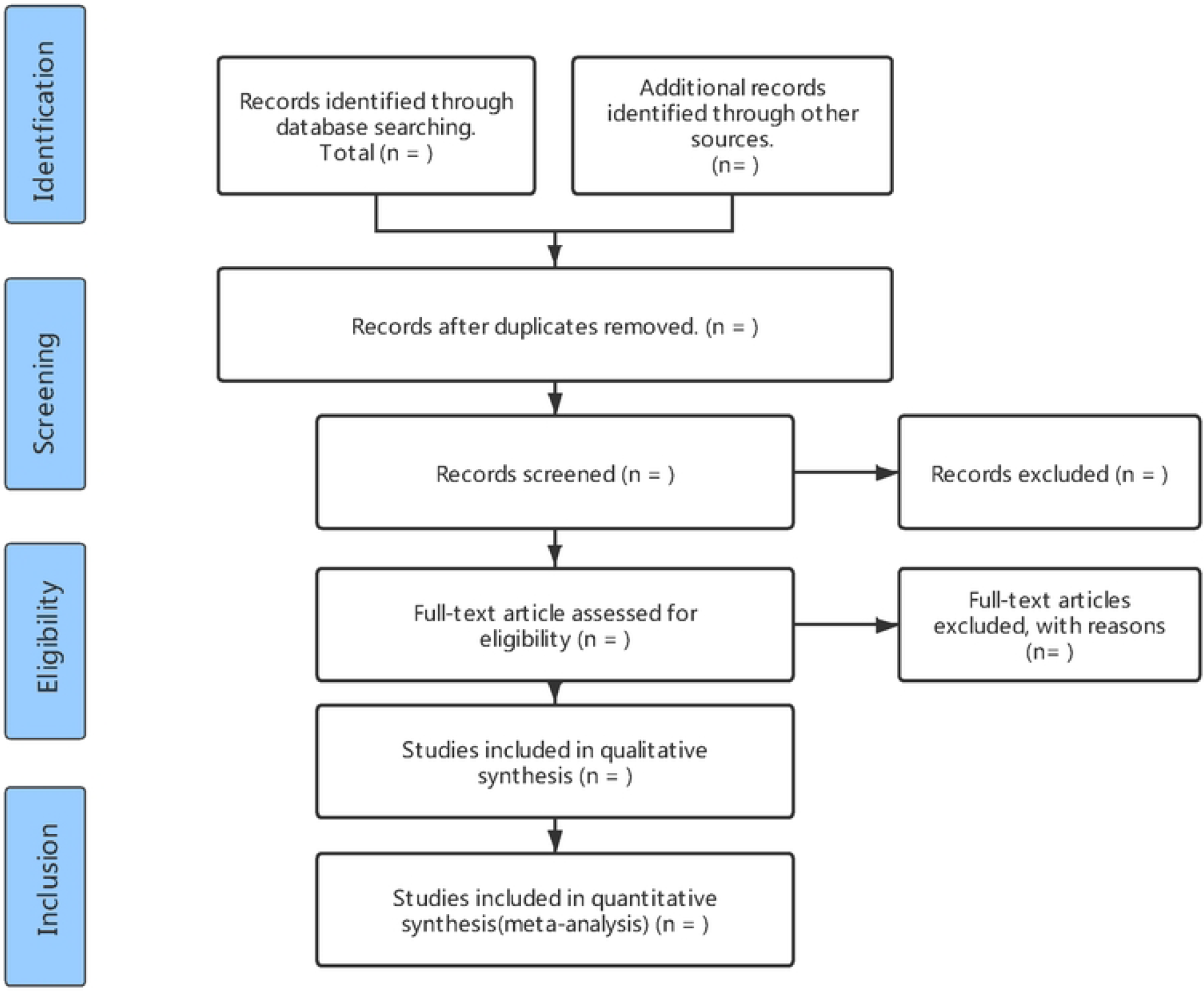
The PRlSMA flow char of study. PRlSMA=Preferred Reporting Items for Systematic reviews and Meta-Analyses.

### 2.8.2 Data extraction

Two independent authors performed the data extraction process, and the following items from each trial were arranged.

1. The basic information of the trials or publications: the title of the article, first author, year, location, and language;
2. Participation baseline of the study. The subgroup condition and statistic distribution of the sample size, age, and sex included the study’s inclusion and exclusion criteria. Interventions in the observation group and the control group. Main outcome(s) and addition outcome(s), and their scale basis. Besides, we will contact the lead author if there is any missing or unavailable information.

#### 2.8.3 Risk of bias assessment

We will assess the methodological quality of randomized controlled trials following the RCT quality assessment criteria recommended in the Cochrane Reviewer’s Handbook (5.3). Risk of Bias in Systematic Reviews (ROBIS) tool will evaluate the quality of reviews. Two assessors will check the results independently, and internal discussion will be performed if the results are inconsistent.

#### 2.8.4 Data synthesis

The data will be analyzed and synthesized through Review Manager (v5.3) software. Relative risk (RR) with 95% confidence intervals (CI) will be used for a dichotomous variable and weighted mean difference (WMD), or standard mean difference (SMD) with 95% CI for continuous variable. Cochrane Collaboration will be employed to compute the data synthesis.

#### 2.8.5 Assessment of heterogeneity

I2 statistics will evaluate heterogeneity. If I2 ≥ 50% indicates the possibility of statistical heterogeneity, we will use a random-effects model for the calculation. Otherwise, a fixed-effects model will be used when the heterogeneity is not significant with its I2 < 50%.

#### 2.8.6 Subgroup analysis

When heterogeneity is high, subgroup analyses will be performed for different comparators separately if necessary. We will perform a subgroup analysis when heterogeneity is high. It can be discussed as gender type, treatment duration, and course of the disease.

#### 2.8.7 Sensitivity analysis

The sensitivity analyses will be performed by excluding studies with high risks of bias and outliers that are numerically distant from the rest of the data.

#### 2.8.8 Publication bias

We will make funnel plots to evaluate the potential publication when studies included in the meta-analysis are more than ten. When an asymmetry of the funnel plot implies reporting bias, we will explain possible reasons.

### 2.9 Patient and Public Involvement

Patient and Public Involvement statement are not required since this is a meta-analysis based on published studies.

## 3. Results

The results of this systematic review will be published as a peer-reviewed article.

## 4. Discussion

PD is the most common movement disorder and represents the second most common degenerative disease of the central nervous system [17]. The pathogenesis is still unclear, and the clinical manifestations are complex and diverse, including many motor and non-motor symptoms [18, 19]. Fatigue is one of the most common and most disabling symptoms among patients with PD, and it has a significant impact on their quality of life. Although fatigue has been recognized for a long time, there is less evidence to support any therapeutic approach in PD patients [20, 21]. Acupuncture is an important CAM treatment modality and a substantial part of Traditional Chinese Medicine (TCM)[22]. Many studies have reported that acupuncture effectively relieves fatigue with various diseases [23-25]. However, the specific mechanism of acupuncture in treating fatigue in PD has not been clarified. Therefore, we will use systematic review and meta-analysis to evaluate the effectiveness, safety, and cost-effectiveness of acupuncture in alleviating fatigue and improving function in PD patients, in order to provide more options for treating fatigue in PD and encourage more people to pay attention to acupuncture.

### Study Limitations

At present, the final conclusion of this systematic review may not be complete due to the relatively few studies on the mechanism of acupuncture, the limited number of studies, as well as the limited number of participants included. Moreover, the interpretation of statistical analysis might be difficult due to the numerous methods of acupuncture, the different methods of practitioners and the variety of acupoint prescriptions. It is possible that further studies on larger groups and the rule of acupuncture in the treatment of PD was deeply summarized will be essential to form direct conclusions.

## 5. Conclusions

We hope that this study will provide relevant information on how acupuncture treat fatigue in PD, and this conclusion will provide a reliable basis for the clinical application of acupuncture in PD. The findings of this review may further clarify the mechanism of acupuncture in the treatment of PD, so as to better improve the symptoms of PD patients and promote the rehabilitation of PD patients.

## Data Availability

No datasets were generated or analysed during the current study. All relevant data from this study will be made available upon study completion.

## Abbreviations

PD: Parkinson’s disease
RCT: randomized controlled trials
CNKI: China National Knowledge Infrastructure
VIP: China Science and Technology Journal database
GRADE: Grading of Recommendations Assessment, Development, and Evaluation.

## Author Contributions

L.Y. and Z.J. contributed equally to this work. Conceptualization, L.Y. and L.T.; methodology, L.Y. and W.X.; software, L.Y. and Z.B.; validation, W.X.; formal analysis, L.X., L.T. and Z.J.; investigation, Z.B.; resources, L.T.; data curation, L.X., L.T. and Z.J.; writing—original draft preparation, L.Y.; writing—review and editing, Z.J.; supervision, W.X. and L.T. All authors have read and agreed to the published version of the manuscript.

## Institutional Review Board Statement

Not applicable.

## Informed Consent Statement

Not applicable.

## Data Availability Statement

The dataset generated during the current study is available from the corresponding authors on reasonable request.

